# A Decade of Progress in Artificial Intelligence for Fundus Image-Based Diabetic Retinopathy Screening (2014–2024): A Bibliometric Analysis

**DOI:** 10.1101/2024.11.02.24316635

**Authors:** Yuting Huang, Yunfei Qi, Chi Liu, Fengshi Jing, Changjin Li, Minghao Wang, Congcong Zhu, Peng Gui, Zongyuan Ge, Xiaotong Han

**Author notes:** Co-first authors (Yuting Huang, Yunfei Qi). **Synopsis/Precis:** A comprehensive bibliometric analysis of AI in fundus image-based diabetic retinopathy screening (2014-2024) reveals significant growth in research output, with India, China, and USA leading publications, showing evolution from traditional methods to deep learning approaches.

## Abstract

**Background/Aims:** Diabetic retinopathy (DR) screening using artificial intelligence (AI) has evolved significantly over the past decade. This study aimed to analyze research trends, developments, and patterns in AI-based fundus image DR screening from 2014 to 2024 through bibliometric analysis.

**Methods:** The study analyzed 1,172 publications from the Web of Science Core Collection database using CiteSpace and Microsoft Excel. The analysis included publication trends, citation patterns, institutional collaborations, and keyword emergence analysis.

**Results:** Publications showed consistent growth from 2014-2022, with a peak in 2021. India (26%), China (20.05%), and USA (9.98%) dominated research output. IEEE ACCESS was the leading publication venue with 44 articles. Research evolved from traditional image processing to deep learning approaches, with recent emphasis on multimodal AI models. The analysis identified three distinct phases: CNN-based systems (2014-2020), Vision Transformers and innovative learning paradigms (2020-2022), and large foundation models (2022-2024).

**Conclusion:** The field shows mature development in traditional AI approaches while transitioning toward multimodal learning technologies. Future directions indicate increased focus on telemedicine integration, innovative AI algorithms, and real-world implementation.

**What is already known on this topic:**

- AI-based DR screening has been developing since the 1960s, with significant acceleration after 2014 due to deep learning advances.
- Traditional manual analysis of fundus images is time-consuming and error-prone.

**What this study adds:**

- Comprehensive mapping of research evolution in AI-based DR screening over the past decade.
- Identification of research concentration in specific geographical areas and emerging trends in multimodal AI approaches.

**How this study might affect research, practice or policy:**

- Highlights the need for increased international collaboration and technology sharing.
- Suggests focus areas for future research, including multimodal learning and real-world implementation.
- Provides direction for healthcare organizations and researchers in adopting and developing AI-based DR screening technologies.

## 1 Introduction

Diabetic retinopathy (DR) represents one of the most prevalent chronic complications among diabetic patients and is a leading cause of blindness [1]. As the number of individuals with diabetes continues to grow worldwide, DR has emerged as a significant threat to the quality of life of those affected [2]. Early screening of Diabetic Retinopathy is crucial for preventing vision loss, enabling timely and cost-effective treatment, improving quality of life, and reducing the overall burden on healthcare systems by detecting and managing the condition before it progresses to more severe stages [3], [4].

A common approach for diabetic retinopathy (DR) screening is fundus imaging due to its clear biomarkers, cost-effectiveness, and non-invasive nature [5]. However, traditional manual analysis of fundus images is often slow, prone to errors, and labor-intensive, which limits its use in large-scale and rapid DR screening programs [6]. To address these challenges, the adoption of artificial intelligence (AI)-based automation techniques for screening has become increasingly prevalent [7], [8].

The integration of artificial intelligence (AI) with fundus imaging has a research history that dated back to the 1960s [9]. From then, for nearly half a century until 2014, AI techniques for fundus images primarily relied on image processing algorithms to measure and evaluate the pathological biomarkers present in the images [9], [10]. A significant transition occurred around 2012 with the emergence of Deep Neural Networks (DNN), which heralded the advent of the deep learning era [11]. In contrast to conventional image processing methods, deep learning demonstrates enhanced capabilities in feature representation and pattern recognition, leading to substantial improvements in screening accuracy and efficiency. As a result, from 2014 onward, this technology has been rapidly and widely adopted in the realm, as documented in literature [12].

To date, in retrospect of the past decade since 2014, the field of fundus image-based diabetic retinopathy (DR) screening has undergone revolutionary changes [13], [14]. This evolution can be roughly divided into three stages from a technical perspective. In the first six years until 2020, research focused primarily on convolutional neural network (CNN)-based deep learning systems and their real-world deployment [15], [16]. From 2020 to 2022, new deep learning models, such as Vision Transformers and their variants, emerged alongside a broader adoption of innovative learning paradigms such as self-supervised, unsupervised, and federated learning [17]. In the most recent two years, large foundation models, including large language and vision models, have sparked significant advancements, complemented by techniques like multi-modal learning and generative AI [18]. In a nutshell, the past decade has seen a remarkable evolution of AI in fundus image-based DR screening, with the trend continuing to progress.

The fuel behind this unstoppable trend originates from the substantial efforts of numerous researchers dedicated to integrating novel AI techniques with fundus image-based diabetic retinopathy screening, resulting in a considerable body of academic output. The objective of this paper is to conduct a comprehensive and timely review of the decades’ research progress in this area from 2014 to 2024. This will be achieved by utilizing bibliometric methods to visualize and analyze the relevant literature. The bibliometric analysis will employ a range of metrics, including keywords, the number of publications, high-frequency citations, and contributions from research organizations and countries. This approach will facilitate the identification of research dynamics, future trends, and potential barriers in this field. It is our hope that this study will provide valuable references and insights for researchers and clinicians in related fields.

## 2 Materials and Methods

### 2.1 Initial data collection

Research literature on fundus image-based AI for diabetic retinopathy (DR) screening was searched in the Web of Science (WOS) Core Collection database. The search formula used was “(Fundus OR Retinal) AND (Image OR Imaging OR photo OR photography) AND (Learning OR Network OR model OR intelligence OR CNN OR AI OR Language) AND (Screening OR Diagnosis OR Grading OR Classification) AND (DR OR Diabetic Retinopathy).” The literature types included Article, Proceeding Paper, Review Article, and Early Access, with the publication indexing period set from January 1, 2014, to July 14, 2024, the date this study was conducted. A total of 2,914 documents were retrieved from literature in the initial collection.

### 2.2 The 2^nd^ round inclusion and exclusion criteria

The inclusion criteria for this study were: studies designed to screen for unique DR-related features including microaneurysms, spot and imprint hemorrhages, hard exudates and/or cotton wool spots, vein beading, intraretinal microvascular anomalies (IRMA), and diabetic macular oedema, even if the selected search terms did not appear in their titles or abstracts. The exclusion criteria for this study were (1) Duplicate publications; (2) Informationally Incomplete Literature; (3) Research of Fundus Image-based AI applied to other diseases. The entire data collection workflow is illustrated in Figure 1. Following the inclusion and exclusion criteria, a total of 1,172 documents were eventually selected for bibliometric analysis.

**Figure 1.**
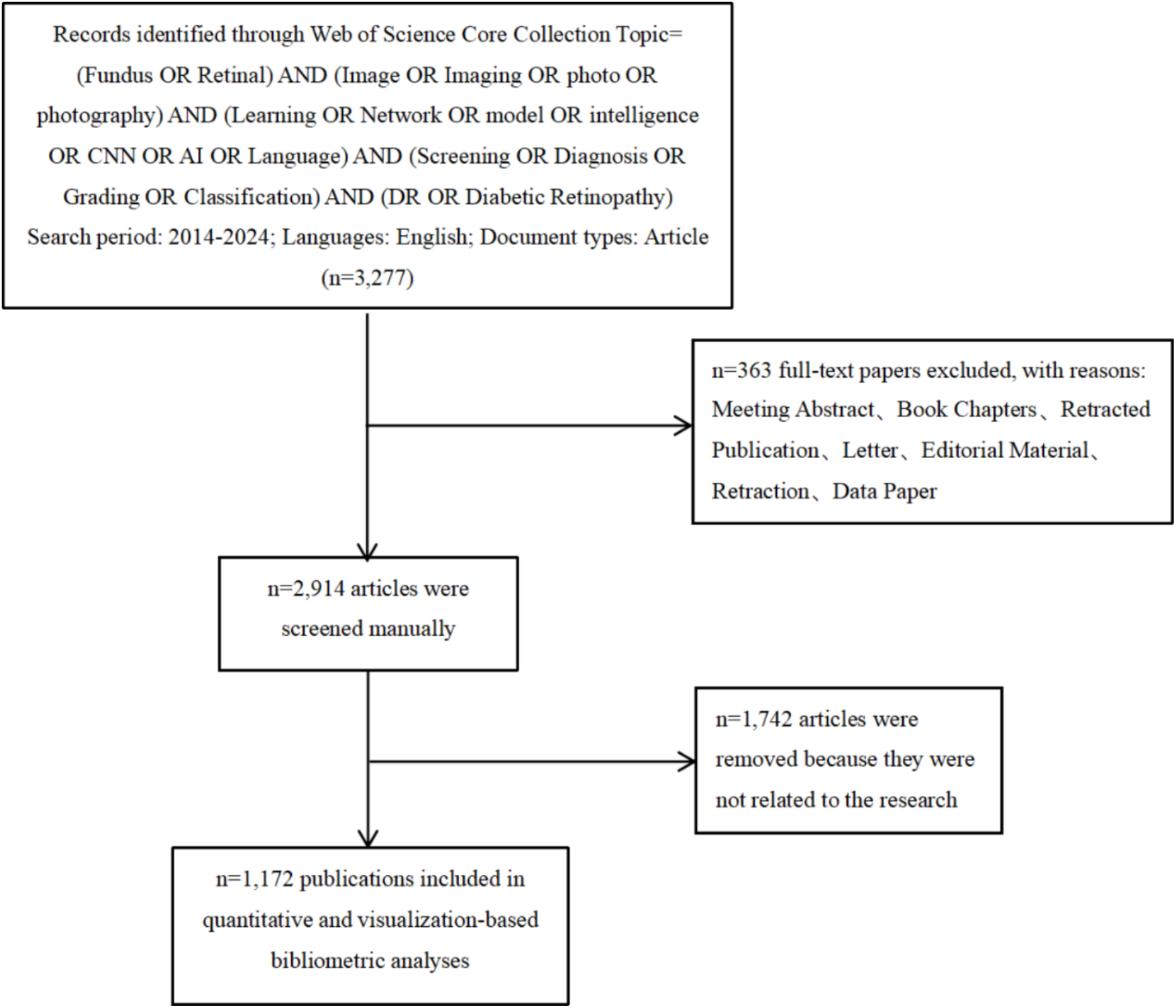
The data collection workflow of this study.

### 2.3 Literature analysis methods

CiteSpace software (Version 6.4.R1) [19] was utilized to analyze the collaborative networks of countries and regions in the selected papers, conduct hotspot analysis of organizations, and perform emergence analysis of keywords. The emergence analysis reveals the frequency of keywords in articles published over a brief period; longer emergence durations indicate sustained keyword relevance and stronger research frontiers. Microsoft Excel (Version 2409) [20] was employed for statistical analysis of publication distribution in the searched literature, the venues of publication, the ranking of highly cited works, and the analysis of organizations and countries.

## 3 Results

### 3.1 Annual distribution of publications

Figure 2 shows a consistent upward trend in the number of articles published by year in the field of fundus image-based AI for diabetic retinopathy (DR) screening. From 2014 to 2016, publications were relatively low, with an average annual growth rate of 10.19%, marking the beginning stage where most studies conducted pilot studies to preliminarily validate deep learning algorithms. From 2017 to 2022, the average annual growth rate surged to 30.58%, with a peak in 2021, which saw an increase of 42 articles from the previous year. However, growth has decelerated since 2023, and data from the first half of 2024 indicates 105 articles published. If this trend persists, growth momentum is likely to continually decline, indicating a saturated equilibrium in this research area.

**Figure 2.**
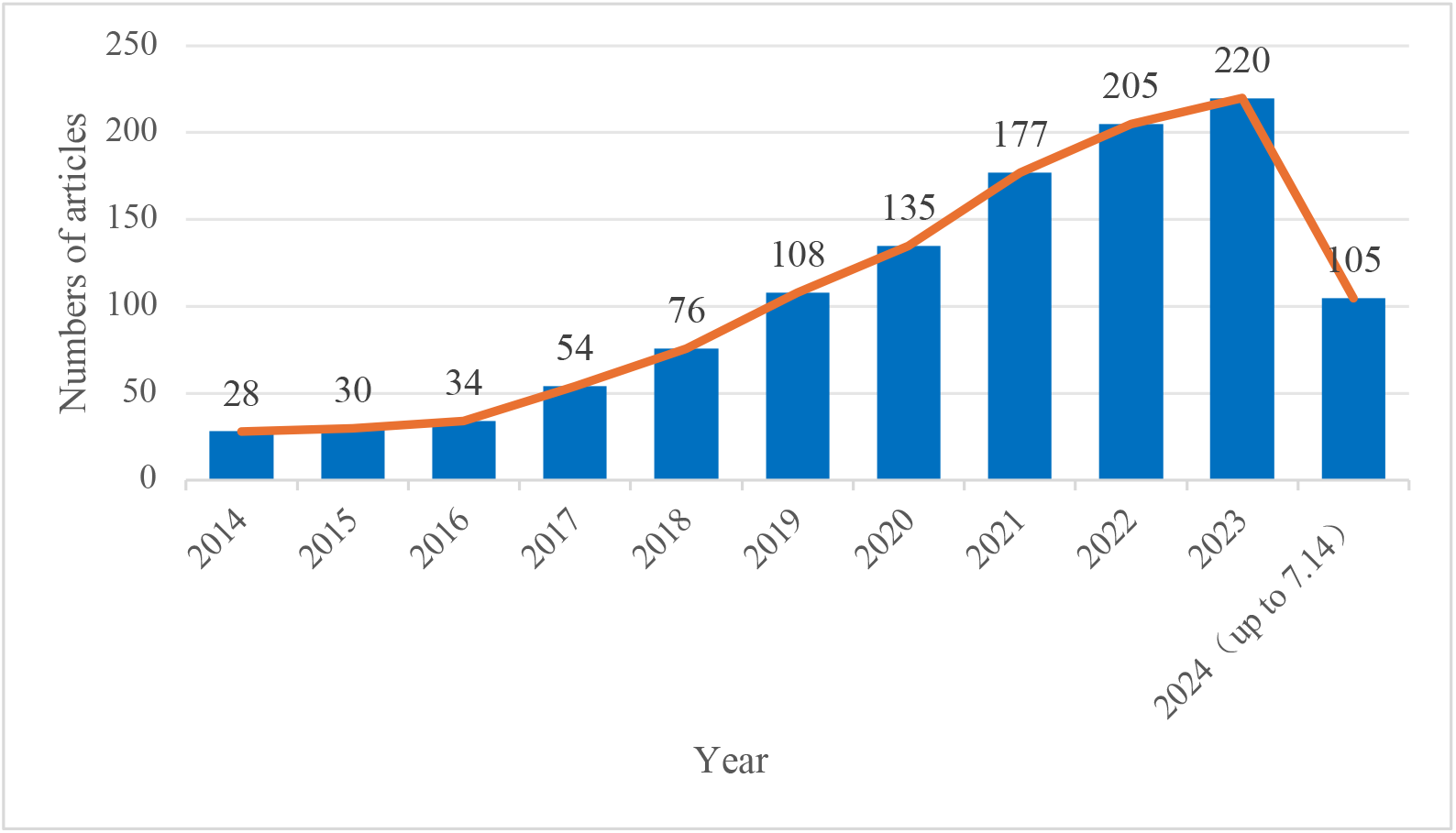
Annual distribution of papers published from 2014 to 2024 on artificial intelligence for fundus image-based DR screening.

### 3.2 Publication Source Analysis

Table 1 analyzes the top 20 journals with the highest number of publications in the reviewed literature. The leading source is IEEE ACCESS, with 44 articles, followed by MULTIMEDIA TOOLS AND APPLICATIONS with 42 articles, and BIOMEDICAL SIGNAL PROCESSING AND CONTROL with 35 articles. The majority of these journals are classified in the Q1 division of the Journal Citation Reports (JCR) by Clarivate, indicating a high academic standard in their fields. Regarding the Journal Impact Factor (JIF), most journals are above 2.5, and COMPUTERS IN BIOLOGY AND MEDICINE has the highest JIF of 7, signifying that its articles are widely cited and recognized in the academic community.

**Table 1.** The top 20 sources with the most publications on artificial intelligence for fundus image-based DR screening.

| Sources | Number of articles | Primary JCR discipline | JCR Category | JIF (2023-2024) |
| --- | --- | --- | --- | --- |
| IEEE ACCESS | 44 | Computer Science | Q2 | 3.4 |
| MULTIMEDIA TOOLS AND APPLICATIONS | 42 | Computer Science | Q2 | 3 |
| BIOMEDICAL SIGNAL PROCESSING AND CONTROL | 35 | Interdiscipline | Q1 | 4.9 |
| DIAGNOSTICS | 21 | Medicine | Q1 | 3 |
| COMPUTERS IN BIOLOGY AND MEDICINE | 19 | Interdiscipline | Q1 | 7 |
| SCIENTIFIC REPORTS | 19 | Interdiscipline | Q1 | 3.8 |
| CMC-COMPUTERS MATERIALS & CONTINUA | 14 | Computer Science | Q3 | 2 |
| SENSORS | 14 | Interdiscipline | Q2 | 3.4 |
| INTERNATIONAL JOURNAL OF IMAGING SYSTEMS AND TECHNOLOGY | 13 | Engineering | Q2 | 3 |
| APPLIED SCIENCES-BASEL | 12 | Interdiscipline | Q1 | 2.5 |
| COMPUTER METHODS AND PROGRAMS IN BIOMEDICINE | 12 | Computer Science | Q1 | 4.9 |
| COMPUTER METHODS IN BIOMECHANICS AND BIOMEDICAL ENGINEERING-IMAGING AND VISUALIZATION | 12 | Engineering | Q4 | 1.3 |
| TRANSLATIONAL VISION SCIENCE & TECHNOLOGY | 12 | Ophthalmology | Q2 | 2.6 |
| EYE | 10 | Ophthalmology | Q1 | 2.8 |
| IEEE JOURNAL OF BIOMEDICAL AND HEALTH INFORMATICS | 10 | Computer Science | Q1 | 6.7 |
| PLOS ONE | 10 | Interdiscipline | Q1 | 2.9 |
| JOURNAL OF INTELLIGENT & FUZZY SYSTEMS | 9 | Computer Science | Q3 | 1.7 |
| MEDICAL & BIOLOGICAL ENGINEERING & COMPUTING | 9 | Computer Science | Q2 | 2.6 |
| ARTIFICIAL INTELLIGENCE IN MEDICINE | 8 | Computer Science | Q1 | 6.1 |
| ELECTRONICS | 8 | Computer Science | Q2 | 2.6 |

Regarding the primary JCR discipline category, among these 20 journals, nine belong to computer science, six belong to the interdisciplinary, two in the engineering, two in the ophthalmology, one in the medicine. This distribution reflects the typical interdisciplinary nature of research in this field. Studies not only focus on methodological innovations of AI techniques in medical practice but also emphasize clinical translation in ophthalmology. The integration of ophthalmic medicine and computer science holds significant potential for advancing this area.

### 3.3 Highly Cited Papers

Table 2 lists the top 10 highly cited papers in this domain from 2014 to 2024. Citation count serves as a key metric to illustrate research trends during this period and the subsequent influence and contributions of the referred work to the community. For example, Varun Gulshan et al. published their article “Development and Validation of a Deep Learning Algorithm for Detection of Diabetic” in 2016 in JAMA [21], a high-impact journal of integrative clinical medicine. The authors developed a deep neural network-based algorithm that can efficiently and accurately automate the detection of diabetic retinopathy and diabetic macular oedema, which has the potential to be applied to improve eye care for diabetic patients. The paper has received widespread attention as a result and has been cited 3,845 times. Another example article “Screening for diabetic retinopathy: new perspectives and challenges,” published by Vujosevic, Stela et al. in 2020 [22], is a highly cited review paper which underscores the impact of novel technologies, including artificial intelligence, telemedicine, and user-friendly imaging devices, on the efficacy, precision, and cost-effectiveness of DR screening.

**Table 2.** The top 10 articles with the most citations on Artificial Intelligence for fundus image-based DR screening.

| Article Title | Year | Authors | Source Title | WoS Core Citation Counts |
| --- | --- | --- | --- | --- |
| Development and Validation of a Deep Learning Algorithm for Detection of Diabetic Retinopathy in Retinal Fundus Photographs | 2016 | Gulshan, V; Peng, L; Coram, M, et al. | JAMA-JOURNAL OF THE AMERICAN MEDICAL ASSOCIATION | 3845 |
| Development and Validation of a Deep Learning System for Diabetic Retinopathy and Related Eye Diseases Using Retinal Images From Multiethnic Populations With Diabetes | 2017 | Ting, DSW; Cheung, CYL; Lim, G, et al. | JAMA-JOURNAL OF THE AMERICAN MEDICAL ASSOCIATION | 1239 |
| Automated Identification of Diabetic Retinopathy Using Deep Learning | 2017 | Gargeya, R; Leng, T | OPHTHALMOLOGY | 737 |
| Improved Automated Detection of Diabetic Retinopathy on a Publicly Available Dataset Through Integration of Deep Learning | 2016 | Abramoff, MD; Lou, YY; Erginay, A, et al. | INVESTIGATIVE OPHTHALMOLOGY & VISUAL SCIENCE | 628 |
| Convolutional Neural Networks for Diabetic Retinopathy | 2016 | Pratt, H; Coenen, F; Broadbent, DM; Harding, SP; Zheng, YL | 20TH CONFERENCE ON MEDICAL IMAGE UNDERSTANDING AND ANALYSIS (MIUA 2016) | 382 |
| Grader Variability and the Importance of Reference Standards for Evaluating Machine Learning Models for Diabetic Retinopathy | 2018 | Krause, J; Gulshan, V; Rahimy, E, et al. | OPHTHALMOLOGY | 288 |
| Screening for diabetic retinopathy: new perspectives and challenges | 2020 | Vujosevic, S; Aldington, SJ; Silva, P, et al. | LANCET DIABETES & ENDOCRINOLOGY | 282 |
| Deep image mining for diabetic retinopathy screening | 2017 | Quellec, G; Charrière, K; Boudi, Y, et al. | MEDICAL IMAGE ANALYSIS | 279 |
| Leveraging uncertainty information from deep neural networks for disease detection | 2017 | Leibig, C; Allken, V; Ayhan, MS, et al. | SCIENTIFIC REPORTS | 258 |
| A Deep Learning Ensemble Approach for Diabetic Retinopathy Detection | 2019 | Qummar, S; Khan, FG; Shah, S, et al. | IEEE ACCESS | 232 |

By analyzing these highly cited articles, we can identify that most of them were published before 2018. Analyzing early highly cited articles is crucial for identifying emerging trends, understanding their impact on subsequent research, and guiding future studies. These articles often introduce foundational concepts and methodologies that shape this research field, establishing benchmarks for quality and credibility. Additionally, they provide historical context, allowing researchers to trace the evolution of ideas and practices over time.

### 3.4 Organizations and Country Regions

A total of 311 institutions from 76 countries or regions participated in the research of the 7,015 pieces of literature under review. The country with the highest publication volume is India, with a total of 307 pieces of published literature, accounting for 26% of the total publication volume. A total of 19% of the publication volume was accounted for by China (Peoples R), with 235 pieces of published literature, representing 20.05% of the total publication volume. The United States (USA) ranked third with 117 pieces of published literature, accounting for 20.05% of the total publication volume. The second largest number of publications is that of China (Peoples R), with 235 publications, accounting for 20.05% of the total number of publications. The third largest number of publications is that of the USA (USA), with 117 publications, accounting for 9.98% of the total number of publications. As shown in Figure 3, the institution with the highest number of publications is the National Institute of Technology (NIT System) in the USA, with a total of 25 publications, representing 2.13 of the total number of publications. The Vellore Institute of Technology (VIT) in India was ranked second with a total of 23 publications, representing 1.96 of the total number of publications. The Egyptian Knowledge Bank (EKB) in Egypt and the University of London in the UK were ranked third with a total of 21 publications, representing 1.79 of the total number of publications.

**Figure 3.**
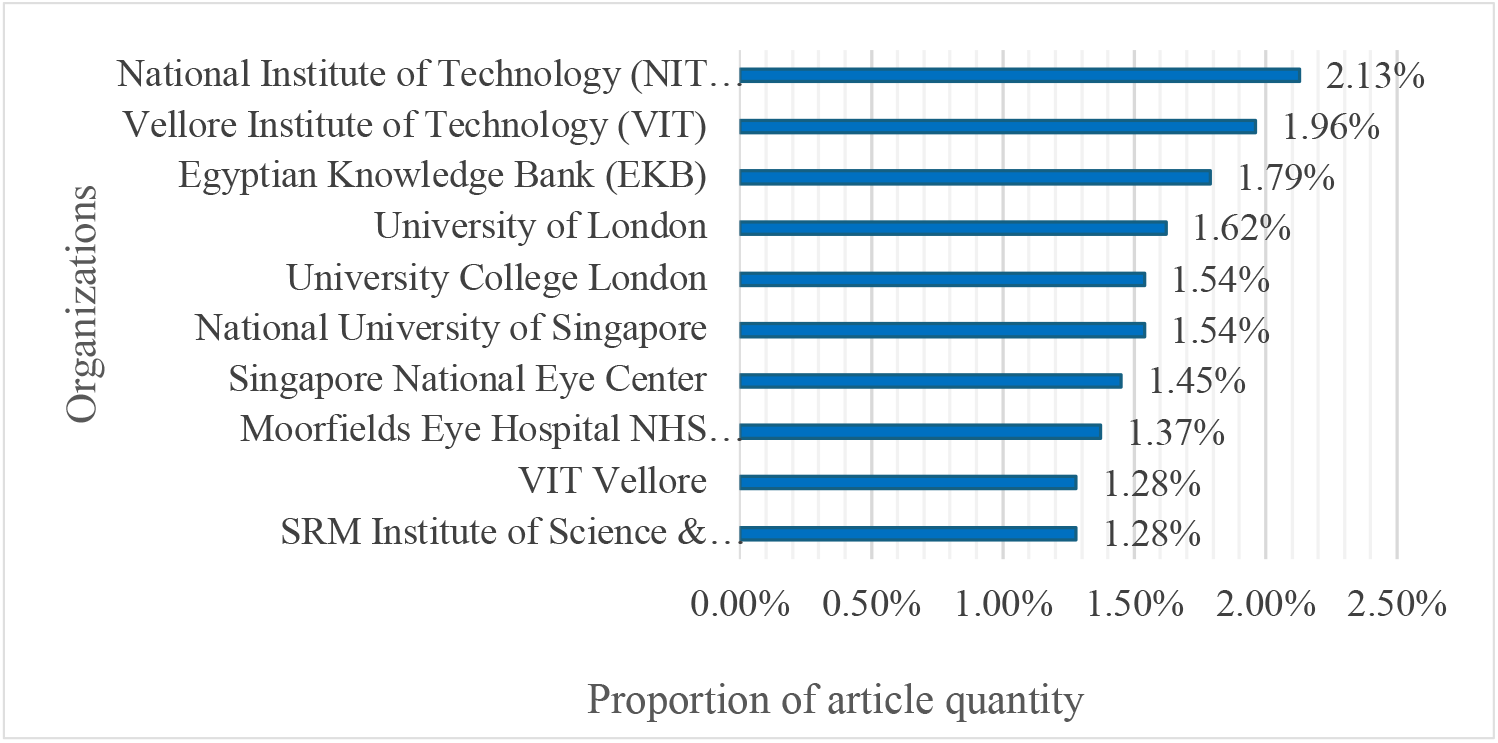
The proportion of articles from the top 10 institutions with the highest number of publications.

It can be found in Figure 4 and Figure 5 that the majority of the top 10 countries and regions in terms of the quantity of related publications are economically developed or undergoing rapid economic development.

**Figure 4.**
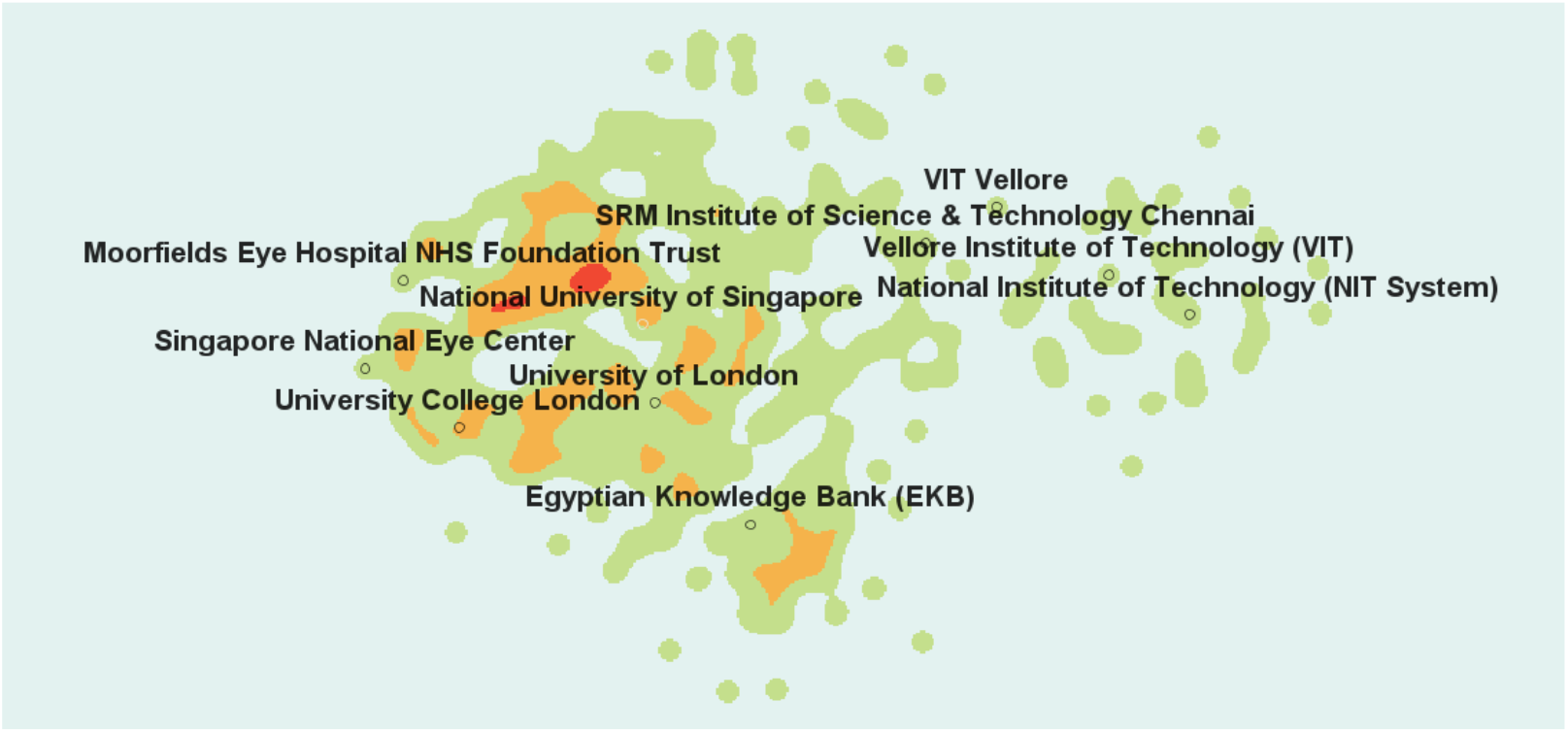
Thermal map of the publishing institution.

**Figure 5.**
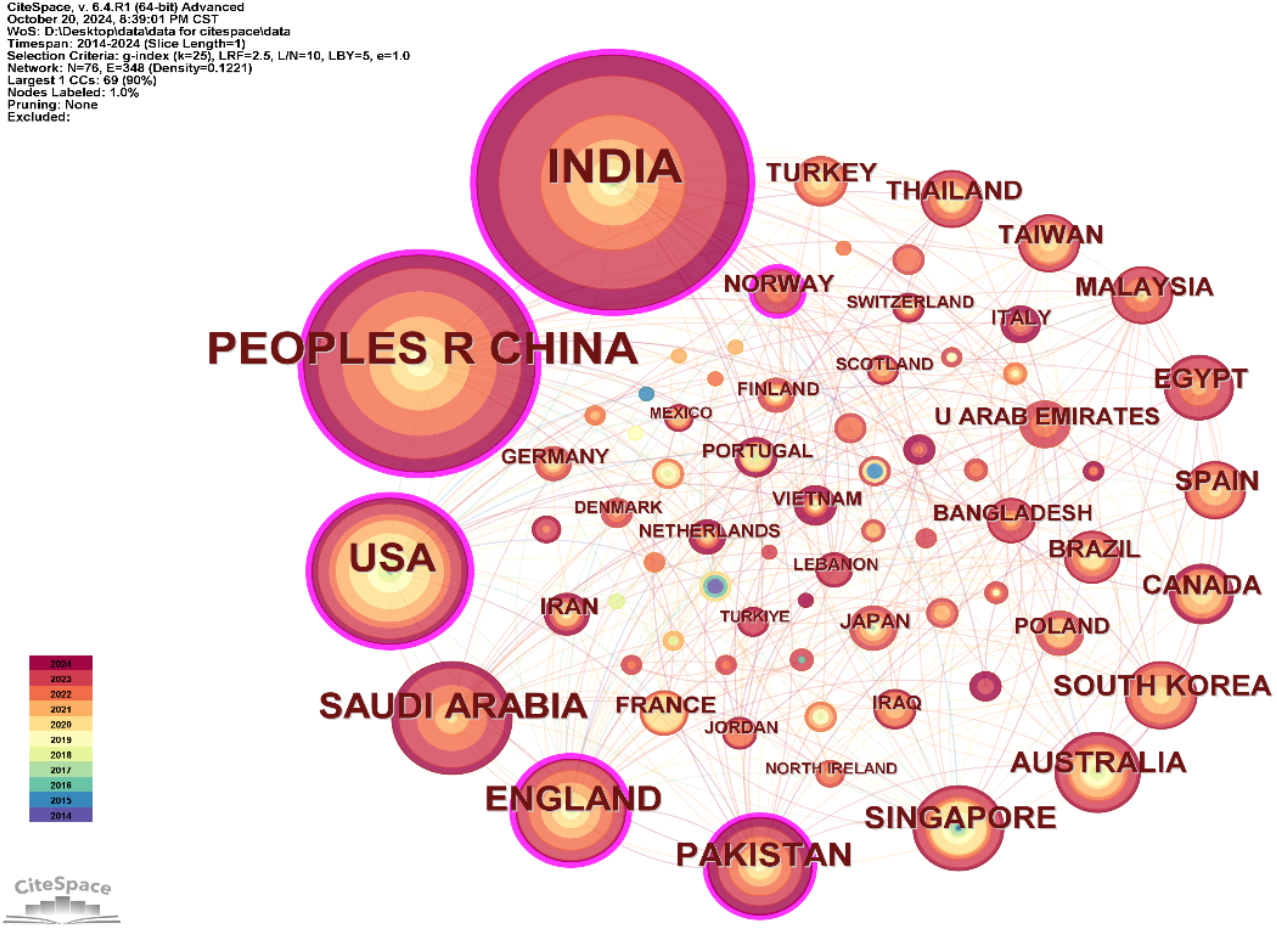
Co-occurrence networks of publishing countries and regions.

In addition, a list of 10 countries with a history of publishing relevant literature for the first time has been provided in Table 3, noting the year of their inaugural publication and the number of publications to date. This list includes several countries from Southeast Asia and Africa. Although these countries have relatively few papers, they show significant potential for growth due to rapidly growing populations, increasing healthcare needs, and weak healthcare foundations. These factors are driving AI technology research and development for fundus imaging in DR screening.

**Table 3.** Ten countries with a history of publishing relevant literature for the first time.

| Country | Year of publication of the first article | Number of articles |
| --- | --- | --- |
| COLOMBIA | 2024 | 1 |
| TUNISIA | 2024 | 1 |
| BAHRAIN | 2023 | 1 |
| LEBANON | 2023 | 6 |
| QATAR | 2023 | 4 |
| TURKIYE | 2023 | 5 |
| UKRAINE | 2023 | 2 |
| MAURITIUS | 2022 | 1 |
| RWANDA | 2022 | 1 |
| SRI LANKA | 2022 | 1 |

### 3.5 Burst terms

Figure 6 is a visual representation of the top 20 keywords with the strongest citation bursts. From 2014 to 2024, several keywords generated significant citation bursts in different years. For example, the term “exudate detection” first appeared in 2014, followed by “retinal image analysis,” “cotton wool spots,” and “blood vessel.” Research activity in the area of fundus image-based AI for DR screening continued into 2019 or 2020. Among the most prominent keywords in 2014 was ‘exudate detection’, which exhibited the highest intensity (3.11) of all keywords. This suggests that it was a subject of particular interest at an early stage.

**Figure 6.**
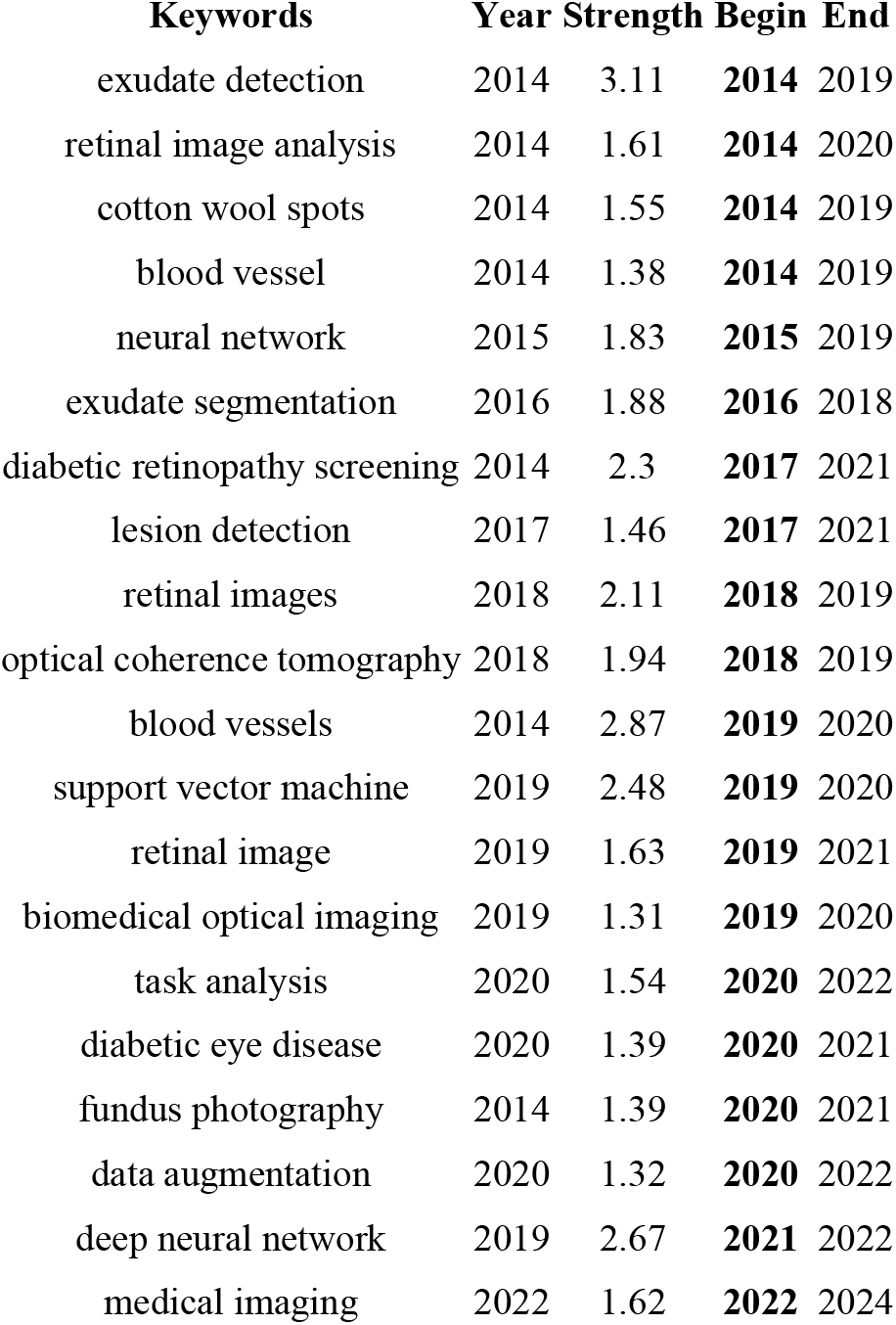
The top 20 keywords with the strongest citation bursts from 2014 to 2024.

The citation burst cycle of keywords also exhibits considerable variation, with the majority of keywords experiencing citation bursts that last between one and two years. This suggests a rapid shift in the research focus. To illustrate, the term ‘exudate segmentation’ was first identified in 2016 and subsequently experienced a notable surge in usage within a relatively brief timeframe. Conversely, the term ‘fundus photography’, which was first documented in 2014, has only recently begun to attract significant attention.

The fluctuations in keywords and their temporal occurrence in the aforementioned graph may serve to illustrate the advancements in technology and the evolution of research focal points within the field. To illustrate, the recently introduced keyword ‘medical imaging’ (2022) is indicative of the potential for continued growth in research activity over the coming years, as evidenced by its initial strength of 1.62 This is a consequence of the development of AI technology. It seems probable that directional image analysis methods (e.g. optical coherence tomography and data enhancement) will continue to grow in the coming years.

As illustrated in the figure, there has been a progressive transition from conventional image analysis approaches (e.g., blood vessel, exudate detection, etc.) to more sophisticated, end-to-end deep learning-based techniques (e.g., ‘neural network’ and ‘deep neural network’). The application of deep learning-related techniques in the field of ophthalmic diabetic retinopathy (DR) has gained increasing attention in recent years, reflecting a broader trend towards the utilization of these artificial intelligence (AI) techniques to facilitate fundus image analysis.

## 4 Discussion

### 4.1 State quo of the research

Our bibliometric analysis examines the intersection of fundus image-based DR screening and AI research from January 2014 to July 2024. A quantitative analysis was performed on various aspects of the dataset. The results show that fundus image AI for DR screening primarily relies on deep learning (DL) algorithms. Currently, external validation of DL models for DR diagnostics is underway in regions with a high volume of publications. Additionally, several AI-based DR screening platforms have received approval and are being marketed by national regulatory authorities in the United States, Europe, Singapore, and China.

The results demonstrate a clear correlation between the number of articles published in the field of AI-based DR screening and the advancement of AI technology. This is illustrated in Figure 1, which depicts the growth of articles in this field over time. The accelerated evolution of deep learning, and the significant advancements in AI for image recognition have led to a notable increase in the number of articles published in this field on an annual basis. Technology in this direction has reached a stage of maturity between 2014 and 2022. Subsequentially, in 2022, the advent of multimodal AI models has prompted a shift in focus within this field towards the development of multimodal learning technology, encompassing not only 2D imaging but also 3D imaging and even texts, as well as multi-modality data processing technology capable of receiving data from disparate channels and sensors. For example, the recent publication in Nature Medicine, titled “Integrated image-based deep learning and language models for primary diabetes care”, developed a multimodal, large-model system, DeepDR-LLM, that integrates image and language [23]. Although only 20 articles have been published in this field as of July 2024, it is anticipated that the volume of research in this direction will continue increasing rapidly.

Our analysis indicates that India, China, and the USA collectively represent over 48.45% of the literature included in the dataset. In light of the considerable population size and the multitude of research organizations present in these three countries, the volume of publications can be considered to be commensurate with the size of the population. The Middle East is disproportionately represented in the dataset, with Saudi Arabia and Pakistan conducting a greater volume of research in this area than the United Kingdom and Australia combined.

### 4.2 Challenges and Opportunities

Since 2014, the number of studies in the field of fundus image-based AI for DR screening has increased rapidly. However, the network of research collaborations in this field is mainly concentrated between top institutions and healthcare organizations. This model of collaboration is closely related to the complexity of AI technology, which requires powerful computational processing power. This remains one of the challenges at present. Concurrently, countries, regions and institutions that have initiated the development of fundus image-based AI for DR screening technology in recent years must reinforce international collaboration and exchanges, as well as prioritize the cultivation of local innovation capabilities. To avoid the formation of information silos, it is essential to proactively adopt and learn from advanced technologies in order to maintain pace with technological advancement and prevent technological lag.

With regard to the field of artificial intelligence, the technology of deep learning is of particular interest, as the majority of current algorithms are based on this approach. In addition to this, a small number of remaining algorithms have also received greater attention, such as those based on binary classification, including support vector machines. In the field of artificial intelligence, different algorithms exhibit distinctive advantages due to their disparate design principles. To enhance the precision of AI-based screening for DR, it is essential to devise effective strategies for integrating these algorithms with imaging techniques. This combination necessitates not only an exhaustive comprehension of the fundamental capabilities of each algorithm, but also the targeted enhancement and optimization of existing algorithms to investigate novel models that are more efficient, accurate and adaptable.

## 5 Conclusion

The accelerated advancement of artificial intelligence (AI) has positioned it as a pivotal driver of modern healthcare advancement. This study identified the research trends in AI for fundus imaging-based DR screening through bibliometric analyses in the past decade. The conclusions include (I) countries with large population bases and developed countries represent the primary focus of DR AI research; (II) future research focus in AI-based DR screening and diagnostic research include telemedicine, innovative AI algorithms, and real-world deployment and evaluation. It is imperative that researchers working in the field of DR must continuously enhance their technical abilities so as to adapt to this transformation. Looking ahead, it is anticipated that AI will become more deeply integrated into the healthcare sector, thereby facilitating more accurate and efficient DR screening, and thus making a greater contribution to global healthcare system.

## Data Availability

All data produced in the present work are contained in the manuscript

## Reference

[1] N. H. Cho et al., ‘IDF Diabetes Atlas: Global estimates of diabetes prevalence for 2017 and projections for 2045’, Diabetes Research and Clinical Practice, vol. 138, pp. 271–281, Apr. 2018, doi: 10.1016/j.diabres.2018.02.023.

[2] J. W. Yau et al., ‘Global Prevalence and Major Risk Factors of Diabetic Retinopathy’, Diabetes Care, vol. 35, no. 3, p. 556, Feb. 2012, doi: 10.2337/dc11-1909.

[3] G. Liew, M. Michaelides, and C. Bunce, ‘A comparison of the causes of blindness certifications in England and Wales in working age adults (16–64 years), 1999–2000 with 2009–2010’, BMJ Open, vol. 4, no. 2, p. e004015, Feb. 2014, doi: 10.1136/bmjopen-2013-004015.

[4] T. Upadhyay, R. Prasad, and S. Mathurkar, ‘A Narrative Review of the Advances in Screening Methods for Diabetic Retinopathy: Enhancing Early Detection and Vision Preservation’, Cureus, vol. 16, no. 2, p. e53586, Feb. 2024, doi: 10.7759/cureus.53586.

[5] B. J. Fenner, R. L. M. Wong, W.-C. Lam, G. S. W. Tan, and G. C. M. Cheung, ‘Advances in Retinal Imaging and Applications in Diabetic Retinopathy Screening: A Review’, Ophthalmol Ther, vol. 7, no. 2, pp. 333–346, Dec. 2018, doi: 10.1007/s40123-018-0153-7.

[6] S. Iqbal, T. M. Khan, K. Naveed, S. S. Naqvi, and S. J. Nawaz, ‘Recent trends and advances in fundus image analysis: A review’, Computers in Biology and Medicine, vol. 151, p. 106277, Dec. 2022, doi: 10.1016/j.compbiomed.2022.106277.

[7] ‘Development and Validation of a Deep Learning System for Diabetic Retinopathy and Related Eye Diseases Using Retinal Images From Multiethnic Populations With Diabetes | Diabetic Retinopathy | JAMA | JAMA Network’. Accessed: Oct. 29, 2024. [Online]. Available: https://jamanetwork.com/journals/jama/fullarticle/2665775

[8] ‘Diagnostic Accuracy of a Device for the Automated Detection of Diabetic Retinopathy in a Primary Care Setting | Diabetes Care | American Diabetes Association’. Accessed: Oct. 29, 2024. [Online]. Available: https://diabetesjournals.org/care/article/42/4/651/36147/Diagnostic-Accuracy-of-a-

[9] Device-for-the-Automated T. Li et al., ‘Applications of deep learning in fundus images: A review’, Med Image Anal, vol. 69, p. 101971, Apr. 2021, doi: 10.1016/j.media.2021.101971.

[10] A. Grzybowski et al., ‘Retina Fundus Photograph-Based Artificial Intelligence Algorithms in Medicine: A Systematic Review’, Ophthalmology and Therapy, vol. 13, no. 8, p. 2125, Jun. 2024, doi: 10.1007/s40123-024-00981-4.

[11] W. Rawat and Z. Wang, ‘Deep Convolutional Neural Networks for Image Classification: A Comprehensive Review’, Neural Computation, vol. 29, no. 9, pp. 2352–2449, Sep. 2017, doi: 10.1162/neco_a_00990.

[12] M. I. Razzak, S. Naz, and A. Zaib, ‘Deep Learning for Medical Image Processing: Overview, Challenges and the Future’, in Classification in BioApps: Automation of Decision Making, N. Dey, A. S. Ashour, and S. Borra, Eds., Cham: Springer International Publishing, 2018, pp. 323–350. doi: 10.1007/978-3-319-65981-7_12.

[13] A. Ikram et al., ‘A Systematic Review on Fundus Image-Based Diabetic Retinopathy Detection and Grading: Current Status and Future Directions’, IEEE Access, vol. 12, pp. 96273–96303, 2024, doi: 10.1109/ACCESS.2024.3427394.

[14] ‘Retinal health screening using artificial intelligence with digital fundus images: A review of the last decade (2012-2023) | IEEE Journals & Magazine | IEEE Xplore’. Accessed: Oct. 29, 2024. [Online]. Available: https://ieeexplore.ieee.org/abstract/document/10713330

[15] ‘A review of convolutional neural networks in computer vision | Artificial Intelligence Review’. Accessed: Oct. 29, 2024. [Online]. Available: https://link.springer.com/article/10.1007/s10462-024-10721-6

[16] ‘Review of deep learning: concepts, CNN architectures, challenges, applications, future directions | Journal of Big Data | Full Text’. Accessed: Oct. 29, 2024. [Online]. Available: https://journalofbigdata.springeropen.com/articles/10.1186/s40537-021-00444-8

[17] A. Younesi, M. Ansari, M. Fazli, A. Ejlali, M. Shafique, and J. Henkel, ‘A Comprehensive Survey of Convolutions in Deep Learning: Applications, Challenges, and Future Trends’, IEEE Access, vol. 12, pp. 41180–41218, 2024, doi: 10.1109/ACCESS.2024.3376441.

[18] A. Bewersdorff et al., ‘Taking the Next Step with Generative Artificial Intelligence: The Transformative Role of Multimodal Large Language Models in Science Education’, Sep. 19, 2024, arXiv: 2401.00832. doi: 10.48550/arXiv.2401.00832.

[19] ‘CiteSpace Home’, CiteSpace. Accessed: Oct. 29, 2024. [Online]. Available: https://citespace.podia.com/

[20] ‘Free Online Spreadsheet Software: Excel | Microsoft 365’. Accessed: Oct. 30, 2024. [Online]. Available: https://www.microsoft.com/en-us/microsoft-365/excel?msockid=192cdc8e90eb6ff63357ce8f91a86e7a

[21] G. V et al., ‘Development and Validation of a Deep Learning Algorithm for Detection of Diabetic Retinopathy in Retinal Fundus Photographs’, JAMA, vol. 316, no. 22, Dec. 2016, doi: 10.1001/jama.2016.17216.

[22] S. Vujosevic et al., ‘Screening for diabetic retinopathy: new perspectives and challenges’, Lancet Diabetes Endocrinol, vol. 8, no. 4, pp. 337–347, Apr. 2020, doi: 10.1016/S2213-8587(19)30411-5.

[23] J. Li et al., ‘Integrated image-based deep learning and language models for primary diabetes care’, Nat Med, vol. 30, no. 10, pp. 2886–2896, Oct. 2024, doi: 10.1038/s41591-024-03139-8.

